# Estimating the distribution of the incubation period of 2019 novel coronavirus (COVID-19) infection between travelers to Hubei, China and non-travelers

**DOI:** 10.1101/2020.02.13.20022822

**Authors:** Char Leung

## Abstract

**Objectives:** Amid the continuing spread of the novel coronavirus (COVID-19), the incubation period of COVID-19 should be regularly re-assessed as more information is available upon the increase in reported cases. The present work estimated the distribution of incubation periods of patients infected in and outside Hubei province of China.

**Methods:** Clinical data were collected from the individual cases reported by the media as they were not fully available on the official pages of the Chinese health authorities. MLE was used to estimate the distributions of the incubation period.

**Results:** It was found that the incubation period of patients with no travel history to Hubei was longer and more volatile.

**Conclusion:** It is recommended that the duration of quarantine should be extended to at least 3 weeks.

## Introduction

An epidemic of viral pneumonia started in Wuhan, the capital of Hubei province in China, in December 2019. A new coronavirus was identified and named by the World Health Organization as COVID-19. It has been found that it is genetically similar to SARS-CoV and MERS-CoV (Zhu et al. 2020). Recently, snakes have been suggested as the natural reservoirs of COVID-19, assuming that the Huanan Seafood Wholesale Market in Wuhan is the origin of the virus (Ji et al. 2020).

As of February 2020, the epidemic of COVID-19 still continued globally and different preventive measures have been implemented by health authorities with quarantine of 14 days being the commonly used. While previous studies have estimated the incubation period of COVID-19 to help determining the length of quarantine, it should be regularly re-assessed as more information is available upon the increase in reported cases.

It has been recently observed that some patients rather had mild symptoms such as cough and low-grade fever or even no symptoms (Chan et al. 2020), constituting greater threat to the effectiveness of entry screening. Against this background, the present work estimated the distribution of incubation periods of patients infected in and outside Hubei. This study found that the incubation period of patients with no travel history to Hubei was longer and more volatile. It is recommended that the duration of quarantine should be extended to at least 3 weeks.

## Materials and methods

Because the details of most cases were reported by the media and were not available on the official web pages of the local health authorities in China, three searches for individual cases reported by the media between 20^th^ January 2020 and 7^th^ February (first cases outside Hubei reported on 20^th^ January 2020) with search terms “pneumonia” AND “Wuhan” AND “age” AND “new” in Chinese were performed on Google from 7^th^, 8^th^, and 9^th^ February. The inclusion of the search term “age” intended to narrow down the search results since the presence of “age” in an article implied a description of an individual case.

Individual cases with time of exposure and symptom onset as well as type of exposure were eligible for inclusion. There was no language restriction. Since most patients did not have complete information about the source of infection, the time of exposure were allowed to be a time interval within which the exposure was believed to lie. The present paper considered two types of exposure, (i) traveling to Hubei, China, and (ii) contact with the source of infection such as an infected person or places where infectious agents stayed. For data accuracy, only confirmed cases outside Hubei province and within China were considered.

The following data were abstracted, (i) location at which the case was confirmed, (ii) gender, (iii) age, (iv) time of exposure, (v) time of first symptom onset, (vi) type of exposure (traveler to Hubei and non-traveler) and (vii) symptoms. Confidence intervals for the average age, proportion of male and different symptoms were constructed. Two-sample t-tests and z-tests for testing the difference in average age and proportions between the traveler and non-traveler groups were also performed.

The incubation period distribution was estimated using maximum likelihood (MLE) that allows interval-censored data (Reich et al. 2009). For an individual case with exact time of exposure and symptom, the likelihood for an exact incubation period observation, *T*, was *f*_*θ*_ (*T*), where *f* and *θ* were the PDF of the incubation period and the set of parameters, respectively. For an individual case with exposure lying between *E*_1_ and *E*_2_, the likelihood for an incubation observation was *F*_*θ*_ (*S* − *E*_1_) − *F*_*θ*_ (*S* − *E*_2_) where *F* and *S* were the CDF of the incubation period and the time of symptom onset, respectively. To find the maximum likelihood estimates of *θ*, the maxima of the sum of the combination of log-likelihood functions were computed with R software.

*T* was assumed to follow lognormal, Weibull and gamma distribution. To ascertain possible difference in distribution between the traveler and non-traveler group, *θ* was adjusted by including additional parameters and indicator variables that took the value 0 or 1 to indicate the type of exposure. For example, the *θ* for a lognormal distribution is given by (*μ* + *Dμ*_*D*_, *σ* + *Dσ*_*D*_)where *D* is the indicator variable.

## Results

A total of 1091 results were generated by Google. A hundred and fifty-two cases from 48 web pages were eligible for inclusion. The cases came from 18 different provinces/municipalities. The temporal profile of all 152 patients is shown in Figure 1. Thirty-nine patients were able to recall the exact date of exposure. Many of these patients stayed in Hubei for a day, or had a friend or family gathering on a particular day. Those could only recall the time interval of exposure largely went to Hubei for sightseeing, work or family visiting, or lived with an infected family member. The earliest history of exposure was 19^th^ December 2019 where a man from Jiangxi went to Wuhan for business procurement whereas the latest history of exposure was 27^th^ January 2020 where a woman from Jiangsu had close contact with an infected person between 22^nd^ and 27^th^ January 2020.

**Figure 1.**
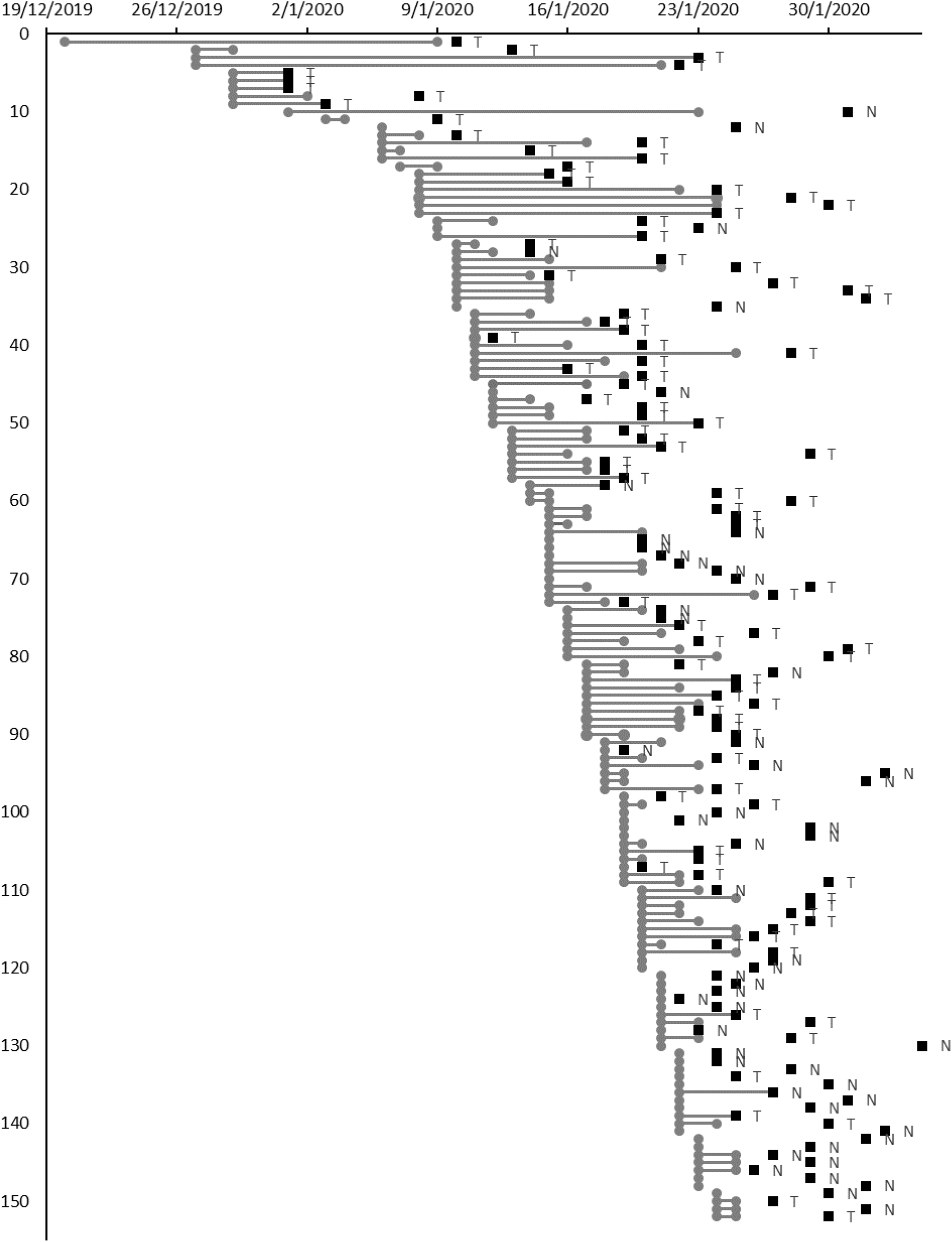
Temporal profile of the 152 patients studied (exposure in grey, symptom onset in black, “T” for traveler to Hubei and “N” for non-traveler)

Of the 151 cases with gender revealed, 93 were male (61.6%, 95% CI 53.3%-69.4%). The average age was 41.2 years (95% CI 38.8-43.5). Travelers to Hubei accounted for 64.5% (95% CI 56.3%-72.1%). With the exception of chill (p = 0.04997), there was no difference in clinical characteristics between the two groups (Table 1). Fever and cough were the most common symptoms regardless of type of exposure.

**Table 1.** Clinical features of patients studied.

|  | All | Travelers to Hubei | Non-travelers | p value |
| --- | --- | --- | --- | --- |
| Cases | 100.0% (152) | 64.5% (98) | 35.5% (54) | 0.000 |
| Age (mean) | 41.2 (145) | 41.1 (94) | 41.2 (51) | 0.959 |
| Gender (male) | 61.6% (151) | 59.8% (97) | 64.8% (54) | 0.543 |
| Symptoms |  |  |  |  |
| Fever | 82.1% (67) | 81.6% (38) | 82.8% (29) | 0.901 |
| Cough | 42.6% (61) | 40.6% (32) | 44.8% (29) | 0.740 |
| General pain | 5.7% (53) | 9.7% (31) | 0.0% (22) | 0.133 |
| Sore throat | 9.3% (54) | 10.0% (30) | 8.3% (24) | 0.834 |
| Weakness | 12.7% (55) | 6.7% (30) | 20.0% (25) | 0.140 |
| Chest pain/discomfort | 7.5% (53) | 10.0% (30) | 4.3% (23) | 0.440 |
| Diarrhoea | 2.0% (51) | 0.0% (29) | 4.5% (22) | 0.246 |
| Chill | 5.7% (53) | 0.0% (29) | 12.5% (24) | 0.04997 |
| Headache | 7.4% (54) | 3.3% (30) | 12.5% (24) | 0.201 |
| Red eye | 1.9% (52) | 0.0% (29) | 4.3% (23) | 0.257 |
| Dyspnea | 7.4% (54) | 9.7% (31) | 4.3% (23) | 0.460 |
| Runny nose and nasal congestion | 2.0% (51) | 0.0% (29) | 4.5% (22) | 0.246 |
| Dizziness | 1.9% (52) | 0.0% (29) | 4.3% (23) | 0.257 |

The results of maximum likelihood estimation are shown in Table 2. The AIC suggested that the Weibull distribution provided the best fit to the data. Both indicator variables of the shape and scale parameters were significant in the Weibull model, suggesting different incubation period distributions between the two groups of patients.

**Table 2.** Results of maximum likelihood estimation.

|  |  | Parameter | Standard error | p value | AIC |  | Mean | Variance |
| --- | --- | --- | --- | --- | --- | --- | --- | --- |
| Log-normal | Location | 0.45 | 0.24 | 0.057 | 207.853 | Traveler to Hubei | 1.7 | 0.8 |
|  | Dispersion | 0.47 | 0.17 | 0.005 |  |  |  |  |
|  | Indicator (location) | 1.27 | 0.26 | 0.000 |  | Non-traveler | 7.1 | 32.8 |
|  | Indication (dispersion) | 0.24 | 0.19 | 0.205 |  |  |  |  |
| Weibull | Shape | 2.30 | 0.90 | 0.011 | 204.643 | Traveler to Hubei | 1.8 | 0.7 |
|  | Scale | 1.99 | 0.46 | 0.000 |  |  |  |  |
|  | Indicator (shape) | -0.57 | 0.93 | 0.543 |  | Non-traveler | 6.9 | 16.7 |
|  | Indicator (scale) | 5.72 | 0.92 | 0.000 |  |  |  |  |
| Gamma | Shape | 4.64 | 3.17 | 0.143 | 204.659 | Traveler to Hubei | 1.8 | 0.7 |
|  | Rate | 2.65 | 1.91 | 0.166 |  |  |  |  |
|  | Indicator (shape) | -2.12 | 3.22 | 0.510 |  | Non-traveler | 6.9 | 18.7 |
|  | Indicator (rate) | -2.28 | 1.91 | 0.233 |  |  |  |  |

## Discussion

The very first observation of the incubation period of COVID-19 came from the National Health Commission of China, reporting the incubation time between 1 and 14 days (NHC 2020). Statistical estimation of the distribution of incubation periods has been done in two other studies (Li et al. 2020, Backer et al. 2020). The present study further explored the difference in incubation periods among different groups of patients. Clinical data were collected from the individual cases reported by the media as they were not fully available on the official pages of the Chinese health authorities. MLE was used to estimate the distributions of the incubation period.

The present work found significant difference in the distribution of the incubation period among travelers to Hubei and non-traveler. The difference was due to both the location and variability, as indicated by the means of 1.8 and 6.9 days and the variances of 0.7 and 16.7 in the Weibull model. Such difference might be due to the difference in infectious dose since travelers to Hubei might be exposed to different sources of infection multiple times during their stay in Hubei. In contrast, patients with no travel history to Hubei were temporarily exposed to their infected relatives, friends or colleagues with mild or even no symptoms.

It is possible that the incubation period of non-travelers was highly volatile, as suggested by the higher variance in the gamma model that provided slightly poorer fit. This could potentially pose a threat to the effectiveness of the existing preventive measures. The duration of quarantine period must be considered with caution.

As a comparison, previous studies on the incubation period for COVID-19 are shown in Table 3. The 95^th^ percentiles reported in previous studies varied between 10.3 and 13.3 days, consistent with the current practice of quarantine period of 2 weeks. However, the present study found that the 95^th^ percentile of non-travelers could be 14.5 days and up to 17.5 days under 95% level of confidence. Coupled with the high variability of the incubation period, it is suggested that the duration of the quarantine period of 3 weeks is deemed more suitable.

**Table 3.** Estimated incubation periods for 2019-nCoV from different studies.

|  | n | Distribution | Mean (days) | 95th percentile (days) |
| --- | --- | --- | --- | --- |
| Li et al. (2020) | 10 | Log-normal | 5.2 (4.1, 7) | 12.5 (9.2, 18) |
| Backer et al. (2020) | 88 | Weibull | 6.4 (5.6, 7.7) | 10.3 (8.6, 14.1) |
| Backer et al. (2020) | 88 | Gamma | 6.5 (5.6, 7.9) | 11.3 (9.1, 15.7) |
| Backer et al. (2020) | 88 | Log-normal | 6.8 (5.7, 8.8) | 13.3 (9.9, 20.5) |
| This study (Travelers to Hubei) | 152 | Weibull | 1.8 (1.0, 2.7) | 3.2 (1.0, 3.8) |
| This study (Non-travelers) | 152 | Weibull | 6.9 (5.5, 8.3) | 14.5 (11.2, 17.5) |

## Data Availability

Data are available in the public domain.

